# Association between cancer and dementia risk in the UK Biobank: evidence of diagnostic bias

**DOI:** 10.1101/2022.10.20.22281285

**Authors:** Jingxuan Wang, Peter Buto, Sarah F. Ackley, Lindsay C. Kobayashi, Rebecca E. Graff, Scott C. Zimmerman, Eleanor Hayes-Larson, Elizabeth Rose Mayeda, Stephen B. Asiimwe, Camilla Calmasini, M. Maria Glymour

## Abstract

**Introduction:** Epidemiological studies have identified an inverse association between cancer and dementia. Underlying methodological biases have been postulated, yet no studies have systematically investigated the potential for each source of bias within a single dataset. We used the UK Biobank to compare estimates for the cancer-dementia association using different analytical specifications designed to sequentially address multiple sources of bias, including competing risk of death, selective survival, confounding bias, and diagnostic bias.

**Methods:** We included 140,959 UK Biobank participants aged ≥55 without dementia before enrollment and with linked primary care data. We used cancer registry data to identify cases of prevalent cancer before UK Biobank enrollment and incident cancer diagnosed after enrollment. We used Cox models to evaluate associations of prevalent and incident cancer with all-cause dementia, Alzheimer’s disease (AD), and vascular dementia. We used time-varying models to evaluate diagnostic bias.

**Results:** The cohort accumulated 3,310 incident dementia diagnoses over a median of 12.3 years of follow-up. All-site incident cancer was positively associated with all-cause dementia incidence (hazard ratio [HR]=1.15, 95% CI: 1.02-1.29), but prevalent cancer was not (HR=1.04, 95% CI: 0.92-1.17). Results were similar for vascular dementia. AD was not associated with prevalent or incident cancer. Dementia diagnosis was substantially elevated in the first year after cancer diagnosis (HR=1.83, 95% CI: 1.42-2.36), after which the association attenuated to null, suggesting diagnostic bias.

**Conclusion:** Health care utilization after a cancer diagnosis may increase chance of a dementia diagnosis, creating potential diagnostic bias in electronic health records-based studies.

## Introduction

Cancer and dementia are leading causes of death globally, and risk of both conditions increase with age.[1–3] Several epidemiological studies have reported an inverse association between cancer and dementia, particularly Alzheimer’s dementia (AD).[4–7] In a recent meta-analysis of 22 cohort and case-control studies, an incident cancer diagnosis was associated with an 11% reduction in the subsequent AD incidence rate.[8] Several biological mechanisms have been proposed to explain why individuals with a history of cancer may have lower dementia incidence rates, including differential cell regulation of senescence and proliferation,[9,10] shared genetic pathways through the *PIN1* and *PARK2* genes,[11,12] and chronic inflammation and immunosenescence.[13]

However, epidemiological results are inconsistent, and several methodological biases have been postulated to explain the findings.[8] In a recent review, Ospina-Romero et al. identified the potential for bias due to competing risk of death, selective survival, confounding, or differential diagnosis.[8] The competing risk of death may introduce bias because people with cancer have a higher mortality risk than cancer-free individuals, and may not live long enough for the diagnosis of dementia.[12,14] Prior evidence using time-varying exposure definitions and evaluating non-fatal cancers suggest this is an unlikely explanation.[7,15] Survival bias may arise if there are unmeasured protective characteristics that promote cancer survival and reduce dementia risk.[13,14] However, a simulation study concluded that selection bias was likely too small to explain the observed inverse association.[16] Confounding may contribute to the inverse relationship in analyses that lacked adequate adjustment for extraneous common causes of cancer and dementia. Diagnostic bias may emerge when cancer diagnoses and treatments cause clinicians to overlook dementia symptoms, delaying the diagnosis of dementia among cancer patients. Simulation studies suggest this can be a particularly influential type of bias.[17] However, diagnostic bias may also work in the opposite direction as cancer patients have more frequent contact with clinicians, increasing their chance of dementia diagnosis.[14]

To better understand what factors may drive the association between cancer and dementia, we used longitudinal data from the UK Biobank to investigate the risks of all-cause dementia, AD, and vascular dementia among survivors of prevalent and incident cancers. We leveraged features of the data to evaluate several potential spurious explanations for a cancer-dementia association. Specifically, we assessed (1) competing risk of death by using transportation injuries as a negative control outcome, [18] (2) survival bias by examining the association between non-melanoma skin cancer (NMSC, a type of cancer that does not substantially increase mortality risk [19]) and dementia, (3) confounding through specification of three sets of covariates, and (4) diagnostic bias by adjusting for the frequency of primary care contact as a proxy for medical surveillance.

## Methods

### Study population

The UK Biobank is a cohort of 502,411 adults aged 40-69 years when recruited from 2006-2010.[20] Participants attended one of 22 assessment centers across England, Wales, and Scotland, where they completed a touchscreen questionnaire and a face-to-face interview with study nurses. Participants also underwent physical measurements and provided blood, urine, and saliva samples. Health outcomes were measured in linked National Health Service (NHS) hospital admissions data, and national death and cancer registers for all participants. Primary care data were available for approximately half of participants. For the current analyses, we included participants who were 55 years or older, free of dementia at the baseline UK Biobank assessment, and had primary care data linked to the UK Biobank database (Figure 1).

**Figure 1:**
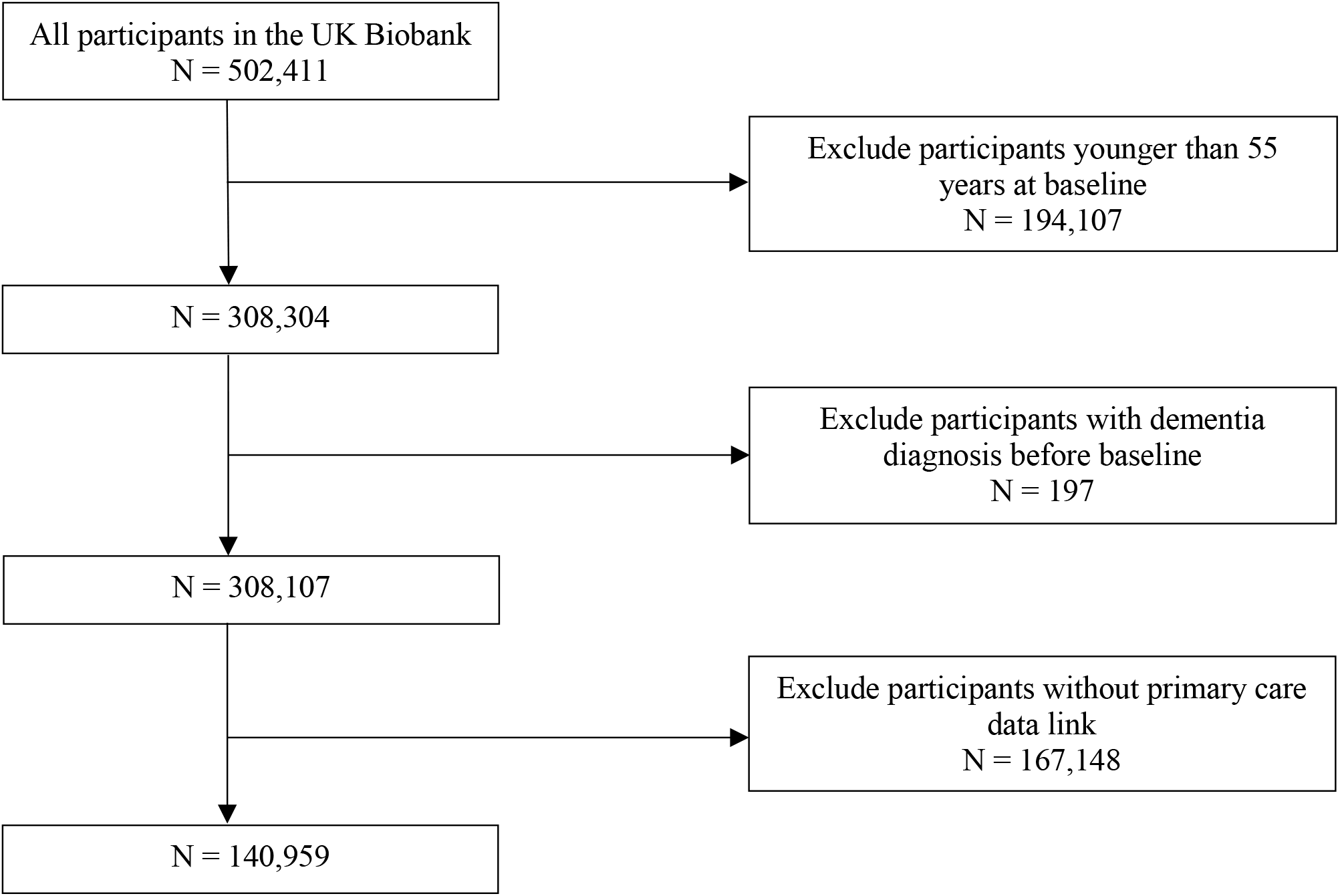
Flow diagram of inclusion criteriap

### Identification of cancer cases

We identified cancer cases and date of diagnosis using data from cancer registries (NHS Digital and Public Health England for England and Wales; NHS Central Register for Scotland). Sensitivity analyses used self-reported cancer diagnoses. In our primary analysis, we included all cancer types except NMSC. We also considered the following common site-specific cancers: NMSC, breast cancer, and prostate cancer, as cancers at each of these sites have been negatively linked to dementia in previous research.[21–23] We treated cancer diagnosis as a time-varying exposure and categorized participants as having (1) prevalent cancer if there was a cancer diagnosis in their medical record prior to the date of UK Biobank enrollment, (2) incident cancer if there was a cancer diagnosis in their medical record *after* their date of UK Biobank enrollment, or (3) no cancer if there was no record of cancer in their medical records, either prior to or after enrollment. All-cause cancer diagnoses were defined as an ICD-10 code of C00-C96 except C44 (NMSC), or an ICD-9 code of 140-209 except 173 (NMSC).

### Ascertainment of dementia cases

Our primary outcome was incident all-cause dementia diagnosis, ascertained via linked NHS hospital admissions, mortality, and primary care data. In the UK Biobank, hospital inpatient data and death records are coded using ICD-9 and ICD-10, and primary care records are coded using Read version 2 (Read v2) and Clinical Terms Version 3 (CTV3), both of which are part of the Systematized Nomenclature of Medicine Clinical Terms (SNOMED-CT).We used a comprehensive list of ICD codes to identify all-cause dementia cases, including AD, vascular dementia, frontotemporal dementia, Lewy body dementia, alcohol-related dementia, and Creutzfeldt-Jakob disease (Supplementary Table 1). Incident dementia was defined as having dementia as a primary or secondary diagnosis in hospital inpatient or primary care data, or a contributory cause of death in mortality data. We studied all-cause dementia as the primary outcome because overlapping and evolving symptoms can lead to ambiguity in the diagnosis of dementia type.[24] In addition, we evaluated the incidence of AD and vascular dementia as secondary outcomes.

### Covariates

The following covariates were included in multivariable analyses: age, sex, race, assessment center, the presence of the APOE-ε4 allele, education, Townsend deprivation index (measure of area-level deprivation), number of clinic visits in the year before study entry, body mass index (BMI), ever smoked, ever used alcohol, physical activity, diabetes, and stroke. Age and sex were acquired from a central registry at recruitment and updated by participants. Race, education, ever smoked, and ever used alcohol were self-reported by participants through a touchscreen questionnaire at baseline. Townsend deprivation index, a composite score measuring socioeconomic status,[25] was calculated prior to baseline based on preceding national census output areas and determined by participants’ postcodes at recruitment. The number of clinic visits in the year before study entry was determined by the number of days the participant had any clinical interactions in the primary care data. BMI was calculated from height and weight that were measured by staff during the baseline visit and treated as a continuous variable. Physical activity was defined as self-report of ≥150 min/week of moderate activity or ≥75 min/week of vigorous activity or an equivalent combination.[26] Diabetes and stroke histories at baseline were identified via linked inpatient, primary care, mortality data, or self-reported medical conditions.

### Statistical analyses

We summarized the baseline characteristics of the study sample stratified by cancer status (no diagnosis, prevalent at baseline, or incident during follow-up). To examine the relationship between a cancer diagnosis and an incident dementia diagnosis, we used Cox proportional hazards regression models. Participants were followed from baseline to the date of first dementia diagnosis, death from any cause, or the latest date at which hospital inpatient data were available (September 30, 2021 for England; July 31, 2021 for Scotland; and February 28, 2018 for Wales), whichever came first. We used age as the time scale because mortality and morbidity risks are expected to increase with age. We conducted all analyses stratified by baseline age categories (55-59, 60-64, 65-69, ≥70 years), to allow the underlying baseline hazard function to vary by age category. Cox proportional hazards regression models were also stratified by assessment center (all models). In our primary analysis, incident cancer was treated as a time-varying covariate: participants with an incident cancer diagnosis contributed person-time to the “no cancer” group until their date of cancer diagnosis, after which they contributed to person-time in the incident cancer group. In addition, we evaluated the aforementioned study biases (Table 2).

#### Competing risk of death

To assess the competing risk of death, we repeated the primary analysis using transportation injuries as a negative control outcome. We chose transportation injuries because there is no known biological mechanism in the relationship between cancer and transportation injuries. In the presence of possible bias from competing risk due to death, mortality due to cancer precludes onset of transportation injuries and we expect to observe a negative association between cancer and transportation injuries.

#### Survival bias

In addition to all cancer sites, we examined the association between NMSC and incident dementia to avoid survival bias because NMSC does not substantially increase mortality risk.

#### Confounding bias

In order to address confounding bias, we sequentially adjusted for three sets of covariates at baseline. We considered potential confounders to be variables that influence both cancer and dementia risk. Model 1 included sex, race, assessment center, and the presence of an APOE-ε4 allele. Model 2 additionally included education, Townsend deprivation index, and the number of clinic visits (as a measure of healthcare utilization). Model 3 additionally included BMI, ever smoked, ever used alcohol, physical activity, diabetes, and stroke. To avoid bias potentially induced by adjusting for a collider,[27] we did not control for factors that were downstream consequences of cancer, for example, cancer treatment or comorbidities known to be cancer sequelae.

#### Diagnostic bias

We adjusted for the frequency of primary care contact in the year before baseline to account for differential medical surveillance among individuals with a history a cancer and individuals with no cancer diagnosis. To evaluate diagnostic bias by cancer status, we additionally calculated the hazard ratios for dementia over five time intervals after the earliest cancer diagnosis (0 – 1 year, >1 – 5 years, >5 – 10 years, >10 – 20 years, and >20 years after cancer diagnosis) compared to individuals with no prior cancer diagnosis. We also assessed the association between past-year health care utilization and incident dementia diagnoses in the primary analysis.

For all Cox models, we assessed violations of the proportional hazards assumption with Schoenfeld residual tests.[28] We repeated the analyses using the secondary outcomes (AD and vascular dementia). In addition, we analyzed site-specific associations between cancer and all-cause dementia for breast and prostate cancer.

We conducted three sensitivity analyses to evaluate robustness of our results. First, we examined the impact of potential incompleteness in the health record data on cancer diagnoses by further including data on self-reported cancer history for all participants.[29] Our second sensitivity analysis assessed the potential impact of missing data using multiple imputation by chained equations with 10 imputations.[30] The percentage of missing variables ranged from 0% for sex, age, assessment center, number of clinic visits, and comorbidities, to 2.7% for self-reported physical activity. Most covariates had less than 1% of values missing. Finally, we repeated the primary analyses restricting to individuals with no prevalent cancer diagnosis at study enrollment and evaluated the association of incident cancer with dementia diagnosis. All statistical analyses were performed using R version 4.0.5.

## Results

### Study Sample Characteristics

Among 140,959 participants who met the inclusion criteria, 12,635 (9.0%) had a recorded cancer diagnosis prior to baseline (referred to as a prevalent cancer), and 17,905 (12.7%) received a first cancer diagnosis during follow-up (referred to as incident cancer). Over a median follow-up of 12.3 years (interquartile range, 11.3-13.1 years), corresponding to 1,671,906 person-years at risk for dementia, there were 3,310 cases of incident all-cause dementia. The analytical sample was predominantly White (96.8%), with 2,321 Asian participants, 904 Black participants, and 1,275 participants who identified as some other race (Table 1). The median baseline age of both prevalent and incident cancer patients was 63.4 years, which was slightly older than cancer-free individuals (62.2 years). Dementia incidence rates increased rapidly with age, regardless of cancer status (Figure 2).

**Table 1:** Baseline characteristics of analytical sample stratified by cancer status

| Characteristic | No cancer<br>No. (%)<sup>a</sup> | Prevalent cancer<br>No. (%)<sup>a</sup> | Incident cancer<br>No. (%)<sup>a</sup> |
| --- | --- | --- | --- |
| Number | 110,419 | 12,635 | 17,905 |
| Median age (IQR) | 62.2 (59.0-65.6) | 63.4 (60.2-66.6) | 63.4 (60.4-66.5) |
| Sex |  |  |  |
| Female | 59,941(54.3) | 8,299 (65.7) | 7,859 (43.9) |
| Male | 50,478(45.7) | 4,336 (34.3) | 10,046 (56.1) |
| Race |  |  |  |
| White | 106,516(96.6) | 12,356 (97.8) | 17,394 (96.9) |
| Black | 718 (0.7) | 69 (0.5) | 117 (0.7) |
| Asian | 2,021 (1.8) | 101 (0.8) | 219 (1.2) |
| Other | 1,021 (0.9) | 102 (0.8) | 152 (0.9) |
| Education |  |  |  |
| Higher | 38,723 (35.1) | 4,277 (33.9) | 5,932 (33.2) |
| Secondary | 35,883 (32.6) | 4,079 (32.3) | 5,570 (31.2) |
| Vocational | 8,018 (7.3) | 857 (6.8) | 1,428 (8.0) |
| Other | 27,610 (25.0) | 3,413 (27.0) | 4,944 (27.7) |
| BMI, mean (SD) | 27.6 (4.7) | 27.6 (4.8) | 29.0 (4.7) |
| Physical activity |  |  |  |
| Yes | 84,143(78.3) | 9,323 (76.1) | 13,500 (77.5) |
| No | 23,315 (21.7) | 2,934 (23.9) | 3,915 (22.5) |
| Ever smoked |  |  |  |
| Yes | 52,041 (47.4) | 6,451 (51.4) | 9,690 (54.5) |
| No | 57,757 (52.6) | 6,107 (48.6) | 8,102 (45.5) |
| Ever used alcohol |  |  |  |
| Yes | 104,978 (95.3) | 11,991 (95.0) | 17,154 (96.1) |
| No | 5,148 (4.7) | 625 (5.0) | 700 (3.9) |
| Townsend deprivation index<sup>b</sup> |  |  |  |
| 1 (least deprived) | 23,221 (21.1) | 2,576 (20.4) | 3,681 (20.6) |
| 2-4 | 68,199 (61.8) | 7,756 (61.4) | 10,984 (61.4) |
| 5 (most deprived) | 18,859 (17.1) | 2,291 (18.2) | 3,226 (18.0) |
| Diabetes |  |  |  |
| Yes | 6,952 (6.3) | 843 (6.7) | 1,385 (7.7) |
| No | 103,467 (93.7) | 11,792 (93.3) | 16,520 (92.3) |
| Stroke |  |  |  |
| Yes | 2,191 (2.0) | 301 (2.4) | 422 (2.4) |
| No | 108,228 (98.0) | 12,334 (97.6) | 17,483 (97.6) |
| Median number of clinic visits<sup>c</sup> (25<sup>th</sup>-75<sup>th</sup> percentile) | 6 (2-11) | 8 (3-14) | 6 (2-12) |
| Median follow-up time (25<sup>th</sup>-75<sup>th</sup> percentile) | 12.3 (11.6-13.1) | 12.2 (11.3-13.0) | 4.9 (2.2-8.2) |

Abbreviations: BMI, body mass index (calculated as weight in kilograms divided by height in meters squared); SD, standard deviation.
<sup>a</sup> Percentages may not sum to 100 due to rounding.
<sup>b</sup> Index quintiles, combining social class, employment, car availability, and housing.
<sup>c</sup> Number of primary care clinics visits in the year before study entry.

**Figure 2:**
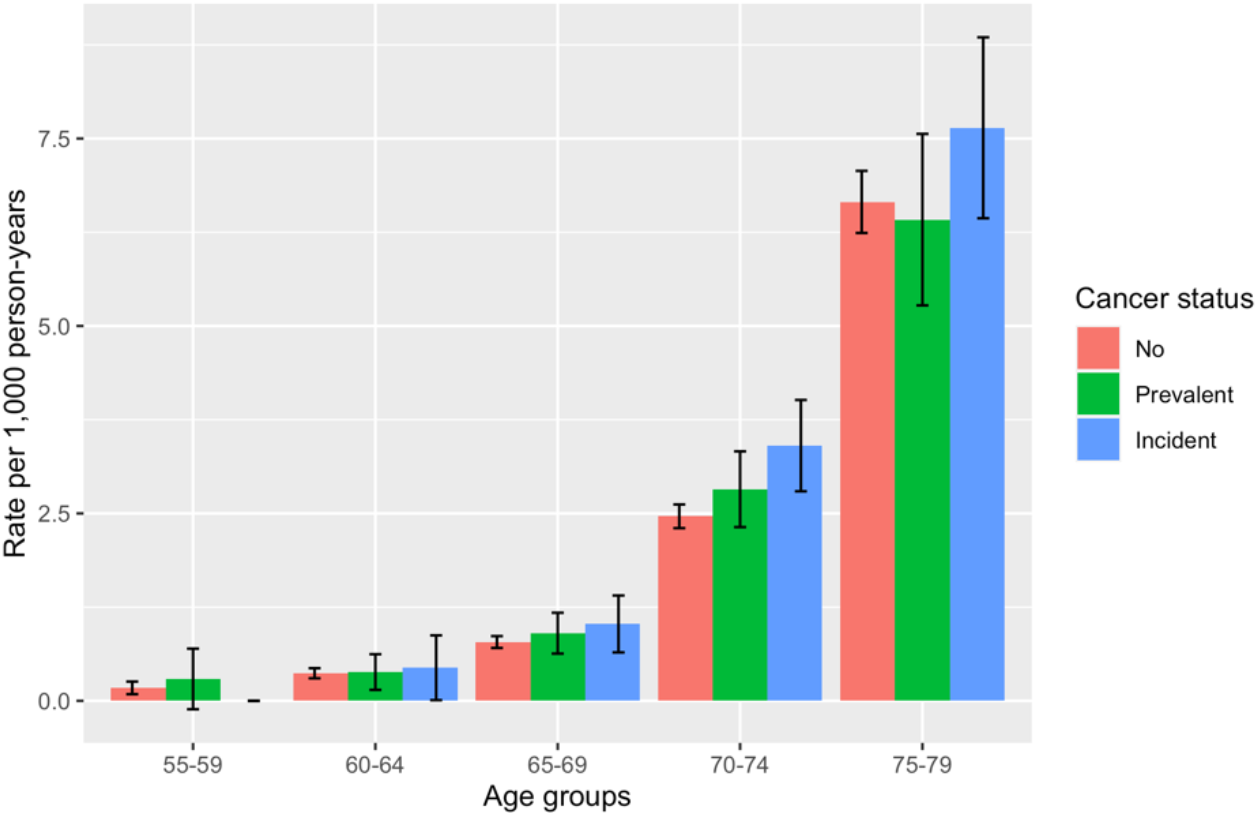
Age-specific all-cause dementia incidence rates. Age-specific incidence rates were calculated by dividing the number of all-cause dementia cases in each age group by the number of person-years of observation in that group. Participants were at risk and contributed person-years from study baseline until the time of first dementia diagnosis, death from any cause, or censoring date. For each age group, a 95% CI for the age-specific incidence rate was calculated assuming a Poisson distribution for the number of cases in that group.

### Associations of history of cancer with dementia

In the minimally adjusted model for the primary analysis (Table 3, Model 1), prevalent cancer was not significantly associated with all-cause dementia (HR = 1.09, 95% CI: 0.97-1.22), while incident cancer was significantly associated with a higher risk of all-cause dementia (HR = 1.16, 95% CI: 1.03-1.30). The fully adjusted model (Table 3, Model 3) yielded similar results (HR = 1.05, 95% CI: 0.93-1.18 for prevalent cancer; HR = 1.15, 95% CI = 1.02-1.29 for incident cancer). A similar hazard ratio is observed for vascular dementia for both prevalent cancer (HR = 1.13, 95% CI: 0.90-1.43) and incident cancer (HR = 1.27, 95% CI: 1.02-1.59; Table 3, Model 3), although not statistically significant for prevalent cancer. Prevalent and incident cancers were not significantly associated with AD, although CIs were wide.

**Table 2:** Types of biases evaluated for alternative explanations

| Bias | Description | Evaluation |
| --- | --- | --- |
| Competing risk of death | People with cancer have a higher mortality risk than cancer-free individuals and may not live long enough for the diagnosis of dementia. | Using transportation injuries as a negative control outcome. |
| Survival bias | There are unmeasured protective characteristics that promote cancer survival and reduce dementia risk. | Examining the association between non-melanoma skin cancer (a type of cancer that does not substantially increase mortality risk) and dementia. |
| Confounding bias | There are common causes of the exposure and outcome that are unaccounted for. | Specifying three sets of covariates. |
| Diagnostic bias | Cancer diagnoses and treatments affect the probability of dementia diagnosis. | Calculating the hazard ratios for dementia over five time intervals after the earliest cancer diagnosis and assessing the association between past-year health care utilization and incident dementia diagnoses. |

**Table 3:** Incident all-cause dementia, AD, and vascular dementia after a cancer diagnosis

| Outcome | No. of events<br>/person-years<sup>a</sup> | Age-<br>adjusted<br>IR<sup>b</sup> | HR | 95% CI | HR | 95% CI | HR | 95% CI |
| --- | --- | --- | --- | --- | --- | --- | --- | --- |
|  |  |  | Model 1<sup>c</sup> |  | Model 2<sup>d</sup> |  | Model 3<sup>e</sup> |  |
| **All-cause dementia** |  |  |  |  |  |  |  |  |
| No cancer | 2,640/1,432,923 | 3.28 | 1.00 | ref. | 1.00 | ref. | 1.00 | ref. |
| Prevalent cancer | 327/143,335 | 3.54 | 1.09 | 0.97-1.22 | 1.04 | 0.92-1.17 | 1.05 | 0.93-1.18 |
| Incident cancer | 343/95,648 | 3.75 | 1.16 | 1.03-1.30 | 1.16 | 1.03-1.30 | 1.15 | 1.02-1.29 |
| **AD dementia** |  |  |  |  |  |  |  |  |
| No cancer | 1,222/1,437,116 | 1.76 | 1.00 | ref. | 1.00 | ref. | 1.00 | ref. |
| Prevalent cancer | 139/143,834 | 1.50 | 0.96 | 0.80-1.14 | 0.93 | 0.77-1.11 | 0.95 | 0.79-1.15 |
| Incident cancer | 147/96,121 | 1.70 | 1.07 | 0.90-1.27 | 1.07 | 0.90-1.27 | 1.05 | 0.87-1.25 |
| **Vascular dementia** |  |  |  |  |  |  |  |  |
| No cancer | 664/1,438,617 | 0.87 | 1.00 | ref. | 1.00 | ref. | 1.00 | ref. |
| Prevalent cancer | 91/144,010 | 0.73 | 1.18 | 0.94-1.48 | 1.08 | 0.86-1.36 | 1.13 | 0.90-1.43 |
| Incident cancer | 95/96,283 | 1.16 | 1.26 | 1.01-1.57 | 1.25 | 1.00-1.56 | 1.27 | 1.02-1.59 |

Abbreviations: AD, Alzheimer's disease; CI, confidence interval; HR, hazard ratio; IR, incidence rate.
<sup>a</sup> Number of dementia cases divided by follow-up time (person-years) in cancer group.
<sup>b</sup> Per 1,000 person-years.
<sup>c</sup> Adjusted for sex, race, and the presence of the APOE-ε4 allele, stratified by assessment center.
<sup>d</sup> Further adjusted for education, Townsend deprivation index, and the number of clinic visits.
<sup>e</sup> Further adjusted for BMI, ever used alcohol, physical activity, diabetes, and stroke, further stratified by ever smoked.

### Assessment of study biases

Incidence of dementia was highest in the first year after cancer diagnosis compared to individuals with no cancer diagnosis (HR = 1.83, 95% CI = 1.42-2.36; Table 4); after the first year post-diagnosis, the association attenuated (HR = 1.03, 95% CI = 0.87-1.23 during 1-5 years after cancer diagnosis; HR = 1.05, 95% CI = 0.89-1.24 during 5-10 years after cancer diagnosis). Similar patterns were observed for AD and vascular dementia. The positive associations between incident cancer and dementia were driven by spike in dementia diagnoses immediately following cancer diagnosis, suggesting diagnostic bias by cancer status. Consistent with this, number of primary care visits per year was associated with higher incidence of all-cause dementia diagnosis (HR = 1.05, 95% CI = 1.04-1.06 for each 3 additional primary care visits per year).

**Table 4:** Incident all-cause dementia, AD, and vascular dementia during five time periods after cancer diagnosis

| Outcome | No. of events<br>/person-years<sup>a</sup> | HR | 95% CI | HR | 95% CI | HR | 95% CI |
| --- | --- | --- | --- | --- | --- | --- | --- |
|  |  | Model 1<sup>b</sup> |  | Model 2<sup>c</sup> |  | Model 3<sup>d</sup> |  |
| **All-cause dementia** |  |  |  |  |  |  |  |
| No cancer | 2,640/1,432,923 | 1.00 | ref. | 1.00 | ref. | 1.00 | ref. |
| 0-1 years | 65/16,960 | 1.83 | 1.42-2.35 | 1.80 | 1.40-2.31 | 1.83 | 1.42-2.36 |
| 1-5 years | 147/60,253 | 1.04 | 0.88-1.24 | 1.03 | 0.87-1.22 | 1.03 | 0.87-1.23 |
| 5-10 years | 163/57,353 | 1.10 | 0.93-1.29 | 1.07 | 0.91-1.26 | 1.05 | 0.89-1.24 |
| 10-20 years | 190/63,426 | 1.10 | 0.95-1.28 | 1.06 | 0.91-1.23 | 1.05 | 0.90-1.23 |
| >20 years | 105/40,991 | 1.06 | 0.87-1.29 | 1.03 | 0.84-1.26 | 1.06 | 0.87-1.31 |
| **AD dementia** |  |  |  |  |  |  |  |
| No cancer | 1,222/1,437,116 | 1.00 | ref. | 1.00 | ref. | 1.00 | ref. |
| 0-1 years | 35/17,018 | 2.22 | 1.57-3.12 | 2.19 | 1.55-3.08 | 2.16 | 1.52-3.08 |
| 1-5 years | 57/60,454 | 0.88 | 0.68-1.16 | 0.88 | 0.67-1.15 | 0.87 | 0.66-1.15 |
| 5-10 years | 64/57,600 | 0.94 | 0.73-1.21 | 0.93 | 0.72-1.19 | 0.91 | 0.70-1.19 |
| 10-20 years | 78/63,710 | 0.91 | 0.72-1.16 | 0.89 | 0.70-1.13 | 0.89 | 0.69-1.13 |
| >20 years | 52/41,172 | 1.06 | 0.80-1.41 | 1.04 | 0.78-1.38 | 1.10 | 0.82-1.46 |
| **Vascular dementia** |  |  |  |  |  |  |  |
| No cancer | 664 /1,438,617 | 1.00 | ref. | 1.00 | ref. | 1.00 | ref. |
| 0-1 years | 17/17,036 | 1.98 | 1.22-3.21 | 1.96 | 1.21-3.18 | 2.10 | 1.29-3.41 |
| 1-5 years | 38/60,554 | 1.10 | 0.79-1.52 | 1.09 | 0.78-1.51 | 1.13 | 0.81-1.57 |
| 5-10 years | 55/57,664 | 1.41 | 1.07-1.87 | 1.38 | 1.04-1.83 | 1.32 | 0.99-1.77 |
| 10-20 years | 43/63,830 | 0.98 | 0.72-1.34 | 0.94 | 0.69-1.29 | 0.99 | 0.72-1.35 |
| >20 years | 33/41,209 | 1.26 | 0.87-1.83 | 1.22 | 0.84-1.78 | 1.28 | 0.87-1.88 |

Abbreviations: AD, Alzheimer's disease; CI, confidence interval; HR, hazard ratio.
<sup>a</sup> Number of dementia cases divided by follow-up time (person-years) in the cancer group.
<sup>b</sup> Adjusted for sex, race, and the presence of the APOE-ε4 allele, stratified by assessment center.
<sup>c</sup> Further adjusted for education, Townsend deprivation index, and the number of clinic visits.
<sup>d</sup> Further adjusted for BMI, ever used alcohol, physical activity, diabetes, and stroke, further stratified by ever smoked.

Supplementary Table 2 shows results for associations of cancer history with transportation injuries. No significant associations between cancer and our negative control outcome, incidence of subsequent transportation injuries were found (HR = 0.96, 95% CI: 0.78-1.18 for prevalent cancer; HR = 0.93, 95% CI: 0.74-1.19 for incident cancer; supplementary Table 2).

Supplementary Table 3 presents results for associations of NMSC with dementia incidence. In the fully adjusted model, incident NMSC diagnosis was associated with lower all-cause dementia incidence (HR = 0.82, 95% CI: 0.67-1.00). The comparison between NMSC and all-site cancer suggested that survival bias may not explain the observed association in the primary analysis. In addition, there were no significant associations between either prevalent or incident prostate cancer diagnoses and all-cause dementia among males (Supplementary Table 3). Similarly, among females, no significant associations between breast cancer and all-cause dementia were found.

In the sensitivity analysis including self-reported cancer diagnoses (Supplementary Table 4), neither prevalent cancer (HR = 1.01, 95% CI: 0.89-1.14) nor incident cancer (HR = 0.97, 95% CI = 0.81-1.17) were significantly associated with incident all-cause dementia. Using multiple imputation to impute missing covariate values (Supplementary Table 5) or excluding individuals with prevalent cancer diagnosis at study enrollment (Supplementary Table 6) did not substantially change our primary results. In addition, there were no significant associations between either prevalent or incident prostate cancer diagnoses and all-cause dementia among males (Supplementary Table 3). Similarly, among females, no significant associations between breast cancer and all-cause dementia were found.

## Discussion

In this large-scale prospective cohort study, we found a significant positive association between incident cancer diagnoses and subsequent all-cause dementia and vascular dementia. These results were driven primarily by a large elevation in the probability of a dementia diagnosis in the year immediately following a new cancer diagnosis. No significant association between overall prevalent or incident cancer diagnoses and AD was found. Incident NMSC diagnosis was associated with a lower risk of all-cause dementia, but prostate and breast cancer were not significantly associated with all-cause dementia.

Prior work suggests a possible inverse biological link between the risk of neoplastic disease and risk of neurodegenerative diagnosis.[31,32] Our findings do not directly support this but evaluate several sources of bias in prior research on this topic. We find little evidence of competing risk of death, selective survival, or confounding bias. We find evidence of a spike in dementia diagnoses immediately following cancer diagnosis, likely due to increased engagement with the health care system and opportunity to receive a dementia diagnosis. These results suggest that diagnostic bias may lead to the observation of positive associations between cancer diagnoses and subsequent diagnoses of dementia received in the course of clinical care. This bias would affect all electronic health records-based studies but would not bias studies in which cognitive outcomes were routinely assessed for all participants as part of the study protocol.

Our results are generally consistent with those from previous studies that ascertained dementia diagnoses from electronic health records and inform interpretation of prior results.[8] Our finding of a temporal trend in the cancer-dementia association suggests that the increased probability of dementia diagnosis in the first year after a cancer diagnosis may be an important source of bias in electronic health records-based studies of cancer and dementia. In Ospina-Romero et al.’s meta-analysis, the pooled HR for studies that ascertained AD from electronic health records was 0.94 (95% CI: 0.58-1.52). For example, a recent population-based cohort study of US veterans that used medical records and claims data thus was also vulnerable to diagnostic bias, found an increased risk of subsequent non-AD dementia among survivors of all cancers (HR = 1.17; 95% CI: 1.15-1.19) and no association between all cancers and AD (HR = 1.00; 95% CI: 0.97-1.03).[33]

On the other hand, in Ospina-Romero et al.’s meta-analysis, community-based studies that did not use medical records to ascertain dementia status and thus classified as less susceptible to AD diagnostic bias showed an inverse association between cancer and AD (HR = 0.73; 95% CI: 0.58-0.90).[8] A study using the Adult Changes in Thought (ACT) cohort reported mixed evidence for the association between all cancers and subsequent dementia among survivors of prevalent cancer before study entry (HR = 0.92; 95% CI: 0.76-1.11) and incident cancer during follow-up (HR = 0.87; 95% CI: 0.64-1.04).[15] In this study, participants were followed up with biennial interviews to identify dementia diagnosis thus differential ascertainment by cancer status is less likely.[34]

Use of electronic health records or other administrative data sources for studying the cancer-dementia link is very appealing because of the large sample sizes available, allowing more precise estimates and investigation of specific cancer and dementia subtypes. Our study confirms a major challenge in such studies, however, due to diagnostic bias. Studies of dementia outcomes based on diagnoses accrued in clinical care must rigorously evaluate and correct for potential diagnostic bias. This potential bias may also be relevant for exposures other than cancer, i.e., any exposure that entails engagement with the health care system (prescriptions, medical treatments, or receipt of care).

### Strengths and limitations

The primary limitations of this study include the limited number of dementia cases, over-representation of healthy and predominantly White individuals with high socioeconomic status (SES) in the UK Biobank, lack of data on time-varying health care utilization, and potential misclassification of dementia diagnoses. Because this cohort is relatively young, with a mean age of 74.4 years (SD=4.3) at the end of follow-up for the age-restricted analytical sample (≥ 55 years at baseline), dementia cases at older ages would not have been observed. Age-specific dementia incidence rates in our sample were substantially lower than in the general UK population.[35] The limited number of incident dementia cases may also have been due to a healthy volunteer selection bias in the UK Biobank.[36] Unlike community-based cohort studies in which routine dementia screening for all participants is completed at predetermined intervals, our study may be vulnerable to diagnostic bias because we derived dementia cases from electronic health records. We attempted to minimize the diagnostic bias by adjusting for the number of primary care clinics visits in the year before study entry. However, we did not consider time-varying health care utilization for individuals with an incident cancer. Finally, comparison between diagnostic codes of all-cause dementia and clinical expert adjudication for a subset of UK Biobank participants showed a positive predictive value of 82.5%,[37] suggesting potential misclassification of dementia diagnosis.

The major strengths of this cohort study include the prospective design, large sample size, long follow-up period, and rich data that allow the assessment of potential methodological biases. Detailed individual information on demographics, behaviors, and comorbidities were collected at baseline, which enabled us to control for multiple potential confounders. Our careful consideration for the potential confounders may preclude the bias induced by inappropriate adjustment for covariates. We also adjusted for the frequency of primary care clinic visits in order to account for potentially differential health care utilization by cancer status. Survival bias due to differential mortality in the cancer and cancer-free groups was mitigated by separating prevalent and incident cancer, and separately investigating NMSC, which does not substantially increase mortality.[19] In addition, our analysis of transportation injuries after a cancer diagnosis did not suggest differential mortality between cancer and cancer-free groups.

### Conclusion and future work

We observed significantly higher incidence of dementia among incident cancer patients than their cancer-free counterparts among adults aged ≥55 in the UK Biobank. Many of these dementia cases were diagnosed in the first year following a cancer diagnosis, suggesting that increased health care utilization following a cancer diagnosis may increase the likelihood of dementia ascertainment. Hence, diagnostic bias could contribute to or explain the association between cancer and dementia observed in electronic health records-based studies. In conclusion, this comprehensive study did not directly support a biologic relationship between cancer and dementia but highlights a bias that may obscure this association in electronic health records-based studies of this topic. Future studies should address these methodological biases to improve causal inference and expand evidence on the etiologies of cancer and dementia. Studies that leverage different methodological approaches, such as evaluating genetic linkage and using neuroimaging biomarkers, may be required to fully understand the relationship between cancer and dementia. Rigorous quantification and correction of diagnostic bias is necessary for electronic health record-based research on the determinants of dementia.

## Data Availability

All data produced are available online at https://www.ukbiobank.ac.uk/

