## Supplementary Tables 1-6 for "Association between cancer and dementia risk in the UK Biobank: evidence of diagnostic bias"

**Supplementary Table 1: ICD codes for dementia**

| Description | Code | Subtype |
|---|---|---|
| **ICD-9** | | |
| Alzheimer’s disease | 331.0 | AD |
| Vascular dementia | 290.4 | VD |
| Frontotemporal dementia | 331.1 | FTD |
| Jakob-creutzfeldt disease | 046.1 | O |
| Alcohol-induced persisting amnestic disorder | 291.1 | O |
| Alcohol-induced persisting dementia | 291.2 | O |
| Senile dementia, uncomplicated | 290.0 | N |
| Presenile dementia | 290.1 | N |
| Senile dementia with delusional or depressive features | 290.2 | N |
| Senile dementia with delirium | 290.3 | N |
| Other specified senile psychotic conditions | 290.8 | N |
| Unspecified senile psychotic condition | 290.9 | N |
| Dementia in conditions classified elsewhere | 294.1 | N |
| Other persistent mental disorders due to conditions classified elsewhere | 294.8 | N |
| Senile degeneration of brain | 331.2 | N |
| **ICD-10** | | |
| Dementia in Alzheimer’s disease | F00 | AD |
| Dementia in Alzheimer’s disease with early onset | F00.0 | AD |
| Dementia in Alzheimer’s disease with late onset | F00.1 | AD |
| Dementia in Alzheimer’s disease, atypical or mixed type | F00.2 | AD |
| Dementia in Alzheimer’s disease, unspecified | F00.9 | AD |
| Alzheimer’s disease | G30 | AD |
| Alzheimer’s disease with early onset | G30.0 | AD |
| Alzheimer’s disease with late onset | G30.1 | AD |
| Other Alzheimer’s disease | G30.8 | AD |
| Alzheimer’s disease unspecified | G30.9 | AD |
| Vascular dementia | F01 | VD |
| Vascular dementia of acute onset | F01.0 | VD |
| Multi-infarct dementia | F01.1 | VD |
| Subcortical vascular dementia | F01.2 | VD |
| Mixed cortical and sub-cortical vascular dementia | F01.3 | VD |
| Other vascular dementia | F01.8 | VD |
| Vascular dementia, unspecified | F01.9 | VD |
| Binswanger’s disease | I67.3 | VD |
| Dementia in Picks disease | F02.0 | FTD |
| Circumscribed brain atrophy | G31.0 | FTD |
| Sporadic Creutzfeldt-Jakob disease | A81.0 | O |

| | | |
|---|---|---|
| Dementia in Creutzfeldt-Jacob disease | F02.1 | O |
| Dementia in Huntington’s disease | F02.2 | O |
| Dementia in Parkinson’s disease | F02.3 | O |
| Dementia in HIV disease | F02.4 | O |
| Mental and behavioural disorders due to use of alcohol – amnesic syndrome | F10.6 | O |
| Dementia in other diseases classified elsewhere | F02 | N |
| Dementia in other specified diseases classified elsewhere | F02.8 | N |
| Unspecified dementia | F03 | N |
| Delirium superimposed on dementia | F05.1 | N |
| Senile degeneration of brain | G31.1 | N |
| Other specified degenerative diseases of nervous system | G31.8 | N |

Abbreviations: AD, Alzheimer’s disease; FTD, frontotemporal dementia; N, no subtype specified; O, other dementia subtype; VD, vascular dementia.

**Supplementary Table 2: Risk of transportation injuries after a cancer diagnosis and during five time periods after a cancer diagnosis**

| Cancer diagnosis | No. of events /person-years[a] | HR | 95% CI | HR | 95% CI | HR | 95% CI |
|---|---|---|---|---|---|---|---|
| | | Model 1[b] | | Model 2[c] | | Model 3[d] | |
| **Cancer status** | | | | | | | |
| No cancer | 1,118/1,416,686 | 1.00 | ref. | 1.00 | ref. | 1.00 | ref. |
| Prevalent cancer | 108/141,810 | 0.99 | 0.81-1.22 | 0.95 | 0.77-1.16 | 0.96 | 0.78-1.18 |
| Incident cancer | 82/94,717 | 0.95 | 0.75-1.20 | 0.93 | 0.73-1.17 | 0.93 | 0.74-1.19 |
| **Time after cancer** | | | | | | | |
| No cancer | 1,118/1,416,686 | 1.00 | ref. | 1.00 | ref. | 1.00 | ref. |
| 0-1 years | 12/16,789 | 0.89 | 0.50-1.57 | 0.88 | 0.50-1.55 | 0.92 | 0.52-1.63 |
| 1-5 years | 41/59,685 | 0.80 | 0.58-1.11 | 0.78 | 0.57-1.08 | 0.80 | 0.58-1.10 |
| 5-10 years | 53/56,883 | 1.09 | 0.82-1.45 | 1.03 | 0.77-1.37 | 1.01 | 0.75-1.36 |
| 10-20 years | 49/62,851 | 0.95 | 0.71-1.27 | 0.91 | 0.68-1.22 | 0.94 | 0.70-1.27 |
| >20 years | 35/40,318 | 1.17 | 0.83-1.65 | 1.14 | 0.81-1.60 | 1.12 | 0.78-1.60 |

Abbreviations: CI, confidence interval; HR, hazard ratio.

[a] Number of dementia cases divided by follow-up time (person-years) in the cancer group.

[b] Adjusted for sex, race, and the presence of the APOE-ε4 allele, stratified by assessment center.

[c] Further adjusted for education, Townsend deprivation index, and the number of clinic visits.

[d] Further adjusted for BMI, ever used alcohol, physical activity, diabetes, and stroke, further stratified by ever smoked.

**Supplementary Table 3: Risk of incident all-cause dementia after diagnosis of non-melanoma skin cancer, breast cancer, or prostate cancer.**

| Exposure | No. of events /person-years[a] | HR | 95% CI | HR | 95% CI | HR | 95% CI |
|---|---|---|---|---|---|---|---|
| | | Model 1[b] | | Model 2[c] | | Model 3[d] | |
| **NMSC** | | | | | | | |
| No cancer | 2,447/1,355,977 | 1.00 | ref. | 1.00 | ref. | 1.00 | ref. |
| Prevalent cancer | 75/34,427 | 0.90 | 0.71-1.13 | 0.88 | 0.70-1.12 | 0.89 | 0.70-1.13 |
| Incident cancer | 118/39,519 | 0.86 | 0.71-1.04 | 0.87 | 0.72-1.05 | 0.82 | 0.67-1.00 |
| **Breast cancer**[e] | | | | | | | |
| No cancer | 1,153/739,010 | 1.00 | ref. | 1.00 | ref. | 1.00 | ref. |
| Prevalent cancer | 60/32,347 | 0.99 | 0.75-1.30 | 0.94 | 0.71-1.24 | 1.02 | 0.77-1.35 |
| Incident cancer | 32/13,488 | 1.13 | 0.79-1.60 | 1.15 | 0.81-1.63 | 1.14 | 0.79-1.65 |
| **Prostate cancer**[f] | | | | | | | |
| No cancer | 1,294/619,967 | 1.00 | ref. | 1.00 | ref. | 1.00 | ref. |
| Prevalent cancer | 22/12,300 | 0.69 | 0.45-1.05 | 0.62 | 0.41-0.95 | 0.68 | 0.44-1.03 |
| Incident cancer | 62/20,171 | 0.82 | 0.63-1.06 | 0.84 | 0.65-1.09 | 0.86 | 0.66-1.11 |

Abbreviations: CI, confidence interval; HR, hazard ratio; NMSC, non-melanoma skin cancer.
[a] Number of dementia cases divided by follow-up time (person-years) in the cancer group.
[b] Adjusted for sex, race, and the presence of the APOE-ε4 allele, stratified by assessment center.
[c] Further adjusted for education, Townsend deprivation index, and the number of clinic visits.
[d] Further adjusted for BMI, ever used alcohol, physical activity, diabetes, and stroke, further stratified by ever smoked.
[e]Among females only.
[f] Among males only.

**Supplementary Table 4: Risk of incident all-cause dementia, AD, and vascular dementia after a cancer diagnosis, including self-reported cancer history**

| Outcome | No. of events /person-years[a] | HR | 95% CI | HR | 95% CI | HR | 95% CI |
|---|---|---|---|---|---|---|---|
| | | Model 1[b] | | Model 2[c] | | Model 3[d] | |
| **All-cause dementia** | | | | | | | |
| No cancer | 2,006/1,157,675 | 1.00 | ref. | 1.00 | ref. | 1.00 | ref. |
| Prevalent cancer | 318/161,072 | 1.05 | 0.93-1.19 | 1.00 | 0.88-1.13 | 1.01 | 0.89-1.14 |
| Incident cancer | 142/56,282 | 1.00 | 0.84-1.19 | 0.99 | 0.83-1.18 | 0.97 | 0.81-1.17 |
| **AD dementia** | | | | | | | |
| No cancer | 889 /1,161,295 | 1.00 | ref. | 1.00 | ref. | 1.00 | ref. |
| Prevalent cancer | 141/161,633 | 1.03 | 0.86-1.23 | 1.00 | 0.84-1.20 | 1.01 | 0.84-1.22 |
| Incident cancer | 64/56,520 | 1.02 | 0.78-1.32 | 1.01 | 0.78-1.31 | 0.98 | 0.75-1.28 |
| **Vascular dementia** | | | | | | | |
| No cancer | 389/1,163,264 | 1.00 | ref. | 1.00 | ref. | 1.00 | ref. |
| Prevalent cancer | 75/161,966 | 1.28 | 0.99-1.64 | 1.18 | 0.91-1.52 | 1.23 | 0.95-1.60 |
| Incident cancer | 40/56,614 | 1.30 | 0.93-1.82 | 1.29 | 0.93-1.80 | 1.31 | 0.94-1.83 |

Abbreviations: AD, Alzheimer's disease; CI, confidence interval; HR, hazard ratio.
[a] Number of dementia cases divided by follow-up time (person-years) among cancer group.
[b] Adjusted for sex, race, and the presence of the APOE-ε4 allele, stratified by assessment center.
[c] Further adjusted for education, Townsend deprivation index, and the number of clinic visits.
[d] Further adjusted for BMI, ever used alcohol, physical activity, diabetes, and stroke, further stratified by ever smoked.

**Supplementary Table 5: Risk of incident all-cause dementia, AD, and vascular dementia after a cancer diagnosis, with multiple imputation to handle missing data**

| Outcome | HR | 95% CI | HR | 95% CI | HR | 95% CI |
|---|---|---|---|---|---|---|
| | Model 1[a] | | Model 2[b] | | Model 3[c] | |
| **All-cause dementia** | | | | | | |
| No cancer | 1.00 | ref. | 1.00 | ref. | 1.00 | ref. |
| Prevalent cancer | 1.09 | 0.97-1.22 | 1.02 | 0.91-1.15 | 1.03 | 0.91-1.15 |
| Incident cancer | 1.16 | 1.03-1.30 | 1.15 | 1.02-1.29 | 1.14 | 1.02-1.28 |
| **AD dementia** | | | | | | |
| No cancer | 1.00 | ref. | 1.00 | ref. | 1.00 | ref. |
| Prevalent cancer | 0.97 | 0.81-1.16 | 0.94 | 0.79-1.12 | 0.94 | 0.79-1.12 |
| Incident cancer | 1.07 | 0.90-1.27 | 1.06 | 0.89-1.26 | 1.06 | 0.90-1.27 |
| **Vascular dementia** | | | | | | |
| No cancer | 1.00 | ref. | 1.00 | ref. | 1.00 | ref. |
| Prevalent cancer | 1.21 | 0.97-1.51 | 1.11 | 0.89-1.38 | 1.14 | 0.92-1.43 |
| Incident cancer | 1.24 | 1.00-1.54 | 1.22 | 0.98-1.52 | 1.22 | 0.98-1.51 |

Abbreviations: AD, Alzheimer's disease; CI, confidence interval; HR, hazard ratio.
[a]Adjusted for sex, race, and the presence of the APOE-ε4 allele, stratified by assessment center.
[b] Further adjusted for education, Townsend deprivation index, and the number of clinic visits.
[c] Further adjusted for BMI, ever used alcohol, physical activity, diabetes, and stroke, further stratified by ever smoked.

**Supplementary Table 6: Risk of incident all-cause dementia, AD, and vascular dementia after a cancer diagnosis, restricting to individuals with no prevalent cancer diagnosis at baseline**

| Outcome | HR | 95% CI | HR | 95% CI | HR | 95% CI |
|---|---|---|---|---|---|---|
| | Model 1[a] | | Model 2[b] | | Model 3[c] | |
| **All-cause dementia** | | | | | | |
| No cancer | 1.00 | ref. | 1.00 | ref. | 1.00 | ref. |
| Incident cancer | 1.15 | 1.03-1.29 | 1.14 | 1.02-1.28 | 1.14 | 1.01-1.28 |
| **AD dementia** | | | | | | |
| No cancer | 1.00 | ref. | 1.00 | ref. | 1.00 | ref. |
| Incident cancer | 1.06 | 0.89-1.26 | 1.06 | 0.89-1.26 | 1.04 | 0.86-1.24 |
| **Vascular dementia** | | | | | | |
| No cancer | 1.00 | ref. | 1.00 | ref. | 1.00 | ref. |
| Incident cancer | 1.26 | 1.01-1.56 | 1.24 | 1.00-1.55 | 1.26 | 1.01-1.58 |

Abbreviations: AD, Alzheimer's disease; CI, confidence interval; HR, hazard ratio.
[a]Adjusted for sex, race, and the presence of the APOE-ε4 allele, stratified by assessment center.
[b]Further adjusted for education, Townsend deprivation index, and the number of clinic visits.
[c]Further adjusted for BMI, ever used alcohol, physical activity, diabetes, and stroke, further stratified by ever smoked.
